# The economic impact of premature mortality in Cabo Verde: 2016 - 2020

**DOI:** 10.1101/2022.11.21.22282604

**Authors:** Ngibo Mubeta Fernandes, Janilza Solange Gomes Silveira Silva, Domingos Veiga Varela, Edna Duarte Lopes, Janice de Jesus Xavier Soares

**Author notes:** Corresponding author: (NMF).

## Abstract

Mortality analysis studies in Cabo Verde are rare and those that are available are limited to short periods of analysis and to specific population groups. Traditionally used mortality data does not quantify the burden resulting from premature mortality. This study focused on the years of potential life lost (YPLL), years of potential productive life lost (YPPLL) and the costs associated with them in Cabo Verde from 2016 to 2020 and aimed to elucidate the patterns of early mortality in the period analyzed.

Mortality data were obtained from the Ministry of Health, Cabo Verde. Deaths that occurred from 2016 to 2020, aged between one (1) and 73 years old, were analyzed by sex, age group, municipality and cause of death. YPLL, YPPLL and cost of productivity lost (CPL) were estimated using life expectancy and the human capital approach.

A total of 6100 deaths were recorded in the sample population and males represented 68.1% (n = 4,154) of the deaths. The total number of deaths verified corresponded to 145,544 YPLL, of which 69.0% (100,389) were attributed to males. There were 4,634 deaths of individuals of working age, and these resulted in 80 965 YPPLL, with males contributing 72.1% (58,403) of the total YPPLL. The total estimated CPL due to premature death was 98,659,153.23 USD with trauma accounting for the highest estimated losses of 21,580,954.42 USD (21.9%), followed by diseases of the circulatory system 18,843,260.42 USD (19.1%), and certain infectious and parasitic diseases accounted for 16,633,842.70 USD (16.9%).

The study demonstrated the social and economic burden of premature mortality. The YPLL, YPPLL and CPL measures can be used to complement measures traditionally used to demonstrate the burden and loss of productivity due to premature mortality and to support resource allocation and public health policies making in Cabo Verde.

## Introduction

Mortality and average life expectancy represent simple measures to assess the health/disease process of a population. National mortality data help to know the characteristics of the deceased and trends in relation to the causes and circumstances of deaths [1]. They also make it possible to compare patterns within and between populations, also serving as a basis for the evaluation and planning of public health programs.

Crude mortality data, however, have important limitations, such as the fact that they do not reflect the age composition of the population, do not report on preventable deaths in certain age groups and do not quantify the burden of premature mortality [2]. To fill this gap, analysts have increasingly resorted to other measures, including the years of potential life lost.

By definition, years of potential life lost (YPLL) represent an estimate of the average time a person would have lived had they not died prematurely. As an impact measure, YPLL seeks to quantify the socioeconomic burden of premature deaths [3].

To quantify economic loss due to early mortality, one can also calculate the years of potential productive life lost (YPPLL) and cost of productivity lost (CPL). The YPPLL use the lower and upper limits of the productive life period as age cut-off and serve as a basis for calculating CPL [4].

Cost of illness studies are considered an essential evaluation technique in the health sector. By measuring and comparing the economic burden of disease on society, these studies can help health decision-makers establish and prioritize health policies and interventions [5].

Despite significant socioeconomic and health improvements, Cabo Verde still has a high burden of mortality in the 15 to 49 age group [6]. Knowing that this age group falls within the productive life span in Cabo Verde (15 to 65 years) it is extremely important to quantify and understand premature mortality in the archipelago.

Mortality analysis studies in Cabo Verde are scarce and those found are limited to short periods of analysis and to specific population groups [7,8]. Through a rapid literature review, no studies were identified evaluating YPLL and/or costs associated with premature mortality in the archipelago. This study sought to fill this gap, calculating and analyzing the years of potential life lost (YPLL), by municipality, sex and cause of death in Cabo Verde in the period between 2016 and 2020, as well as the costs associated with early mortality in the country.

The results of the study intend to elucidate the patterns of early mortality in the analyzed period, being able to subsidize public policies, mainly at the sanitary level. This work focused on the calculation of years of potential life lost, years of potential productive life lost and the costs associated with these in Cabo Verde from 2016 to 2020.

## Methodology

### General approach

This is an observational, cross-sectional, quantitative and analytical study using secondary data. For data analysis, all deaths that occurred in Cabo Verde from 2016 to 2020, aged between one (1) and 74 years old, were considered. Deaths that occurred in the first year of life were excluded due to the specificity of mortality in this period and due to the social and economic value that this loss represents [9,10].

In this study mortality data were presented by cause of death according to the International Statistical Classification of Diseases and Related Health Problems 10th Revision (ICD-10) [11].

## Data sources and data collection

An anonymized database, provided by the National Health Directorate of the Ministry of Health, containing information collected from death certificates of all deaths that had occurred in Cabo Verde from 2016 to 2020 was used.

Quality control involved several aspects, at different levels. Standards of technical quality requirement for the quality of the database were ensured by two elements of the research team, technicians from the National Institute of Public Health (INSP). The entire process of cleaning and structural analysis of the data (data cleaning) was conducted such as identification of ‘missing data’, systematic errors in jumps and inadequate patterns of filed data.

The statistical analysis of data was performed using the SPSS statistical software (version 26) and Microsoft Excel.

## Estimation methods

Statistical analysis of data started with descriptive statistics, calculating absolute and relative frequencies of deaths that occurred in the period under analysis. Calculations of YPLL, YPPLL and CPL, and their respective absolute and relative frequencies, rates and means were determined. YPLL, YPPLL and CPL were estimated by sex, age group, municipality and cause of death.

The Human Capital Approach was adapted, assuming that an individual would earn a constant value throughout their productive life if it were not interrupted by premature death [12].

YPLL were estimated by adapting the method proposed by Romeder and Mcwhinnie [9]. The maximum age limit was 74 years, the average life expectancy at birth for the Cabo Verdean population in 2019 [13]. The YPLL calculation method for a given factor consisted of the sum of the number of deaths at each age (between 1 and 73 years old) multiplied by the remaining years of life up to 74 years of age. The number of deaths were distributed by age groups of five (5) years. In addition, the midpoint of each age groups was calculated. Subsequently, the midpoint of each age group was subtracted from 74 years, and the difference was multiplied by the number of deaths in each age group. The result represents the estimated number of years of life lost due to a specific cause of death. Thus, the following formula was used to estimate YPLL [4]:

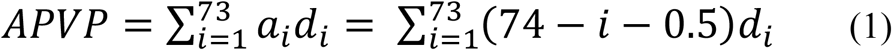

Where: a_i_ – years remaining to reach the age corresponding to the upper limit considered, when death occurs between the ages of i and i + 1 years, using the adjustment of 0.5 assuming that all deaths occurred in the middle of the year and d_i_ – number of deaths between age i and i + 1 years.

To determine the economic losses due to premature deaths during the study period (2016-2020), the YPPLL and CPL method were used [4]:

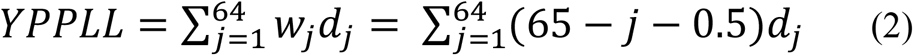

Where: wj – number of years left to complete the age corresponding to the upper limit considered when death occurs between age j and j+1 year; dj – number of deaths between the age of j and j + 1 year, using the adjustment of 0.5 when it is assumed that all deaths occurred in the middle of the year; j – mean age in 5-year age groups for the productive population (15 – 64 years); 65 – retirement age limit in Cabo Verde [14] and 15 – minimum working age[15].

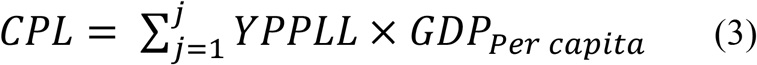

Where: the Gross Domestic Product (GDP) per capita for 2017, of 3 427,49 USD (321,945$00 *Escudos Caboverdianos*) [14] and j as defined previously.

Future values were presented in present values by applying the discount rate of 4.5% practiced by the Bank of Cabo Verde [16]. In addition, a sensitivity analysis was performed applying rates of 3% and 6% to adjust for possible fluctuations in rates [12,17]. To determine the present value of the cost of productivity lost, the following formula was used:

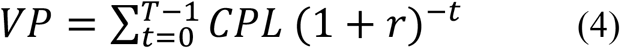

Where: VP – present value, CPL – future value, r – discount rate, t – mean number of years remaining to reach the age corresponding to the upper limit considered in each 5-year age group.

## Ethical considerations

As this study analyzed existing secondary data provided by the National Directorate of Health and published in Annual Health Reports, submission to the National Ethics Committee for Health Research and to the National Commission for Data Protection was not required.

## Results

### Sociodemographic characterization of subjects

#### Sample characterization

The sample consisted of a total of 6100 deaths, aged between 1 and 73 years. The male represented 68.1% (4154) of deaths. Overall, 62.3% (3800) of deaths occurred in the age group between 35 to 64 years. The highest number of deaths occurred in the municipalities of Praia (27.7%; 1692) and São Vicente (17.6%; 1076) Table 1

**Table 1:**
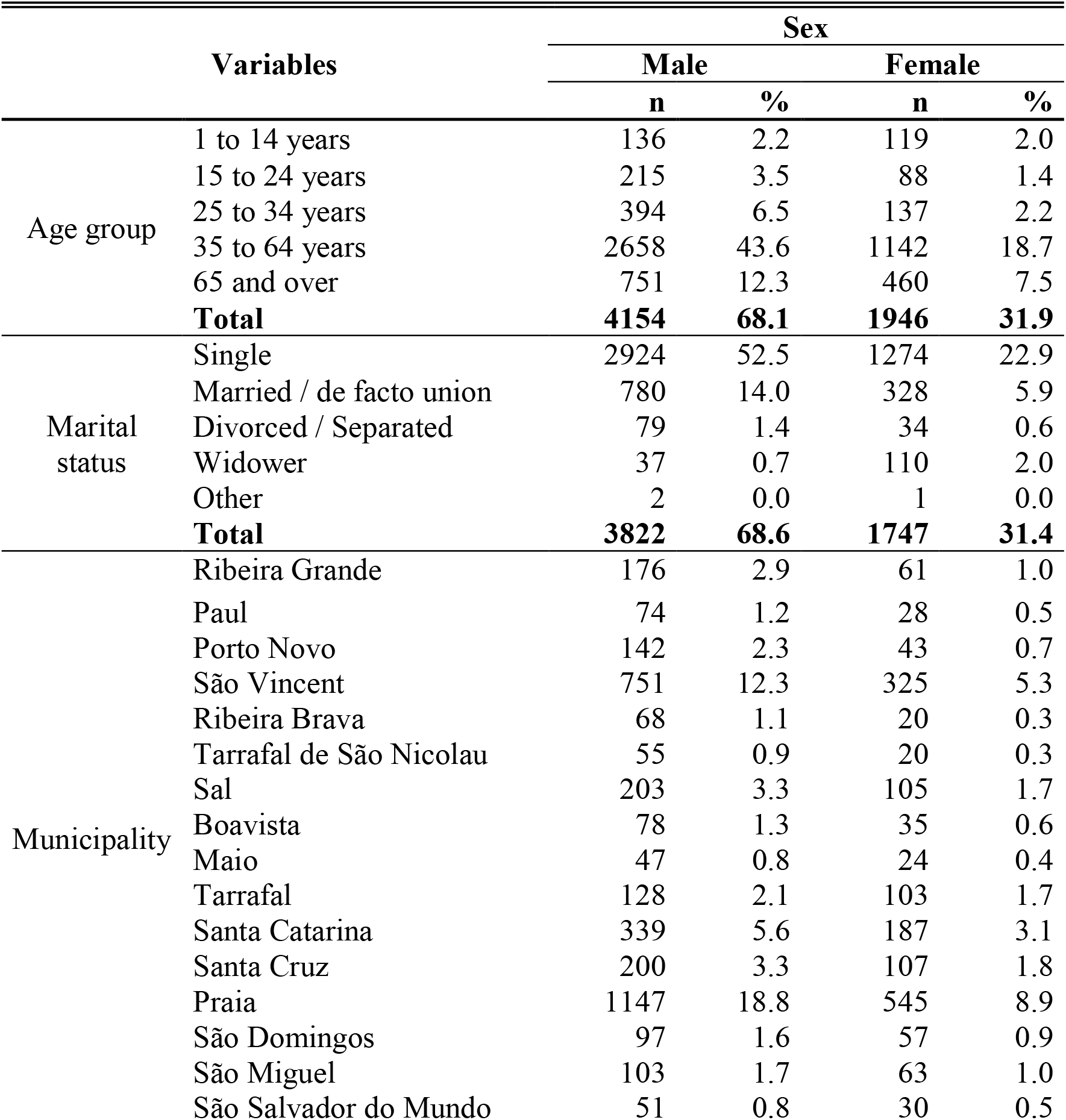
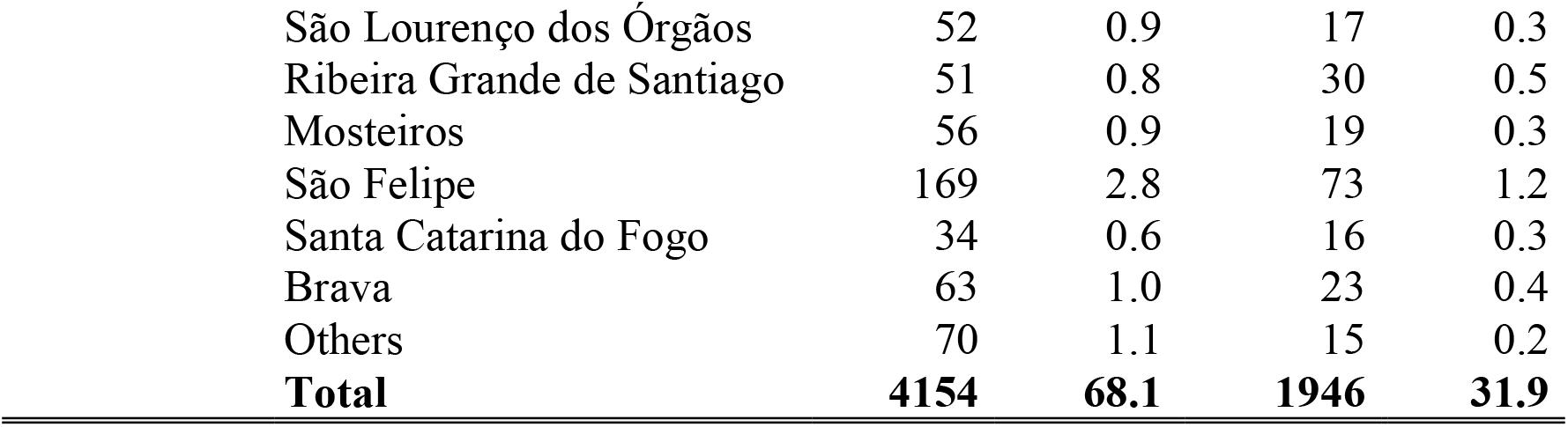
Characterization of the sample population, 2016 to 2020

| Variables |  | Sex |  |  |  |
| --- | --- | --- | --- | --- | --- |
|  |  | Male |  | Female |  |
|  |  | n | % | n | % |
| Age group | 1 to 14 years | 136 | 2.2 | 119 | 2.0 |
|  | 15 to 24 years | 215 | 3.5 | 88 | 1.4 |
|  | 25 to 34 years | 394 | 6.5 | 137 | 2.2 |
|  | 35 to 64 years | 2658 | 43.6 | 1142 | 18.7 |
|  | 65 and over | 751 | 12.3 | 460 | 7.5 |
|  | **Total** | **4154** | **68.1** | **1946** | **31.9** |
| Marital status | Single | 2924 | 52.5 | 1274 | 22.9 |
|  | Married / de facto union | 780 | 14.0 | 328 | 5.9 |
|  | Divorced / Separated | 79 | 1.4 | 34 | 0.6 |
|  | Widower | 37 | 0.7 | 110 | 2.0 |
|  | Other | 2 | 0.0 | 1 | 0.0 |
|  | **Total** | **3822** | **68.6** | **1747** | **31.4** |
| Municipality | Ribeira Grande | 176 | 2.9 | 61 | 1.0 |
|  | Paul | 74 | 1.2 | 28 | 0.5 |
|  | Porto Novo | 142 | 2.3 | 43 | 0.7 |
|  | São Vincent | 751 | 12.3 | 325 | 5.3 |
|  | Ribeira Brava | 68 | 1.1 | 20 | 0.3 |
|  | Tarrafal de São Nicolau | 55 | 0.9 | 20 | 0.3 |
|  | Sal | 203 | 3.3 | 105 | 1.7 |
|  | Boavista | 78 | 1.3 | 35 | 0.6 |
|  | Maio | 47 | 0.8 | 24 | 0.4 |
|  | Tarrafal | 128 | 2.1 | 103 | 1.7 |
|  | Santa Catarina | 339 | 5.6 | 187 | 3.1 |
|  | Santa Cruz | 200 | 3.3 | 107 | 1.8 |
|  | Praia | 1147 | 18.8 | 545 | 8.9 |
|  | São Domingos | 97 | 1.6 | 57 | 0.9 |
|  | São Miguel | 103 | 1.7 | 63 | 1.0 |
|  | São Salvador do Mundo | 51 | 0.8 | 30 | 0.5 |

|  |  |  |  |  |
| --- | --- | --- | --- | --- |
| São Lourenço dos Órgãos | 52 | 0.9 | 17 | 0.3 |
| Ribeira Grande de Santiago | 51 | 0.8 | 30 | 0.5 |
| Mosteiros | 56 | 0.9 | 19 | 0.3 |
| São Felipe | 169 | 2.8 | 73 | 1.2 |
| Santa Catarina do Fogo | 34 | 0.6 | 16 | 0.3 |
| Brava | 63 | 1.0 | 23 | 0.4 |
| Others | 70 | 1.1 | 15 | 0.2 |
| **Total** | **4154** | **68.1** | **1946** | **31.9** |

During the period under analysis, diseases of the circulatory system, infectious and parasitic diseases, neoplasms, diseases of the respiratory system, external causes of morbidity and mortality and injuries, poisonings and certain other consequences of external causes were responsible for 81.7% (5064) of deaths. Diseases of the circulatory system were the main cause of death, accounting for 23.4% (1427) of deaths, followed by some infectious and parasitic diseases (15.4%; 940) and neoplasms (15.3%; 935). It should be noted that in 2020, 112 deaths recorded were due to COVID-19, included in the death group certain infectious and parasitic diseases, Table 2.

**Table 2:** Number of deaths by sex and cause of death, 2016 to 2020.

| Cause of death | Sex |  |  |  |
| --- | --- | --- | --- | --- |
|  | Male |  | Female |  |
|  | n | % | n | % |
| Certain conditions originating in the perinatal period | 2 | 0.03 | 1 | 0.02 |
| External causes of morbidity and mortality | 497 | 8.1 | 72 | 1.2 |
| Diseases of the genitourinary system | 50 | 0.8 | 27 | 0.4 |
| Diseases of the circulatory system | 890 | 14.6 | 537 | 8.8 |
| Diseases of the digestive system | 228 | 3.7 | 88 | 1.4 |
| Diseases of the respiratory system | 435 | 7.1 | 191 | 3.1 |
| Mental and behavioral disorders | 148 | 2.4 | 13 | 0.2 |
| Endocrine, nutritional and metabolic diseases | 106 | 1.7 | 56 | 0.9 |
| Diseases of the nervous system | 83 | 1.4 | 32 | 0.5 |
| Pregnancy, childbirth and puerperium | 0 | 0.0 | 29 | 0.5 |
| Certain infectious and parasitic diseases | 571 | 9.4 | 369 | 6.0 |
| Injury, poisoning and certain other consequences of external causes | 414 | 6.8 | 71 | 1.2 |
| Congenital malformations, deformations and chromosomal abnormalities | 2 | 0.03 | 2 | 0.03 |
| Diseases of the musculoskeletal system and connective tissue | 3 | 0.05 | 1 | 0.02 |
| Diseases of the skin and subcutaneous tissue | 6 | 0.1 | 2 | 0.03 |
| Diseases of the blood and blood-forming organs and certain disorders involving the immune mechanism | 5 | 0.1 | 9 | 0.1 |
| Symptoms, signs and abnormal clinical and laboratory findings, not elsewhere classified | 150 | 2.5 | 75 | 1.2 |
| Neoplasms [tumors] | 564 | 9.2 | 371 | 6.1 |
| **Total** | **4154** | **68.1** | **1946** | **31.9** |

### Estimates of years of potential life lost in Cabo Verde, 2016 to 2020

Of the total number of deaths reported from 2016 to 2020, 42.9% (6100) occurred in the age group from 1 to 73 years, which corresponded to 145,544 YPLL. In the study period, the highest number of YPLL (21,978) were in 2020. The results obtained showed that the total mean YPLL per death in the five-year period varied from 22.6 to 25.0, Table 3.

**Table 3:** Years of potential life lost by sex, Cabo Verde, 2016 to 2020.

| YPLL by sex |  |  |  |  |  |  |  |  |  |  |  |  |
| --- | --- | --- | --- | --- | --- | --- | --- | --- | --- | --- | --- | --- |
| Year | Male |  |  |  | Female |  |  |  | Total |  |  |  |
|  | Deaths | YPLL | YPLL | Mean | Deaths | YPLL | YPLL rate | Mean | Deaths | YPLL | YPLL rate | Mean |
| 2016 | 787 | 19920 | 78.4 | 25.3 | 379 | 9259 | 37.3 | 24.4 | 1166 | 29179 | 58.1 | 25.0 |
| 2017 | 723 | 17095 | 66.2 | 23.6 | 347 | 8552 | 34.0 | 24.6 | 1070 | 25647 | 50.4 | 24.0 |
| 2018 | 868 | 21513 | 82.1 | 24.8 | 425 | 9585 | 37.8 | 22.6 | 1293 | 31098 | 60.3 | 24.1 |
| 2019 | 818 | 19884 | 74.8 | 24.3 | 360 | 8228 | 32.1 | 22.9 | 1178 | 28111 | 53.8 | 23.9 |
| 2020 | 958 | 21978 | 81.5 | 22.9 | 435 | 9532 | 36.8 | 21.9 | 1393 | 31509 | 59.6 | 22.6 |

Results showed that the leading contributors to YPLL between 2016 and 2020 were diseases of the circulatory system, certain infectious and parasitic diseases, external causes of morbidity and mortality and injuries, poisonings and certain other consequences of external causes, which accounted for 71% (103,336) of the total, of YPLL Table 4 In the study period, the highest number of YPLL (31,509) was 2020. In this year, diseases of the circulatory system accounted for 5,913 YPLL, followed by certain infectious and parasitic diseases (5,312), external causes of morbidity and mortality (4,298), neoplasms (3,618), diseases of the respiratory system (3,287) and injuries, poisonings and certain other consequences of external causes (3,077) which together represented 80.9% of YPLL in that year, Table 4.

**Table 4:** Evolution of years of potential life lost by cause of death, 2016 to 2020.

| Cause of death | Reference year |  |  |  |  |
| --- | --- | --- | --- | --- | --- |
|  | 2016 | 2017 | 2018 | 2019 | 2020 |
| Certain conditions originating in the perinatal period | 71 | 0 | 71 | 71 | 0 |
| External causes of morbidity and mortality | 5229 | 3911 | 4020 | 3663 | 4298 |
| Diseases of the genitourinary system | 200 | 247 | 298 | 102 | 248 |
| Diseases of the circulatory system | 4820 | 4343 | 5003 | 4724 | 5913 |
| Diseases of the digestive system | 1523 | 1478 | 1250 | 1568 | 1460 |
| Diseases of the respiratory system | 2317 | 2154 | 3869 | 2848 | 3287 |
| Mental and behavioral disorders | 827 | 896 | 859 | 518 | 920 |
| Endocrine, nutritional and metabolic diseases | 671 | 682 | 691 | 930 | 609 |
| Diseases of the nervous system | 860 | 626 | 855 | 557 | 901 |
| Pregnancy, childbirth and puerperium | 83 | 203 | 156 | 452 | 291 |
| Certain infectious and parasitic diseases | 4697 | 4284 | 5734 | 4256 | 5312 |
| Injury, poisoning and certain other consequences of external causes | 3647 | 3042 | 3483 | 3344 | 3077 |
| Congenital malformations, deformations and chromosomal abnormalities | 71 | 57 | 0 | 138 | 0 |
| Diseases of the musculoskeletal system and connective tissue | 0 | 52 | 73 | 0 | 12 |
| Diseases of the skin and subcutaneous tissue | 22 | 43 | 7 | 2 | 26 |
| Diseases of the blood and blood-forming organs and certain disorders involving the immune mechanism | 0 | 42 | 169 | 171 | 60 |
| Symptoms, signs and abnormal clinical and laboratory findings, not elsewhere classified | 833 | 760 | 1258 | 1295 | 1483 |
| Neoplasms [tumors] | 3311 | 2831 | 3306 | 3476 | 3618 |
| **Total** | **29179** | **25647** | **31098** | **28111** | **31509** |

Males accounted for 69.0% (100,389) of the total estimated YPLL and the total mean loss per death was 23.9 for both sexes, 24.2 for males and 23.2 for females. The highest proportion of YPLL for females (22.1%; 9984) was attributable to certain infectious and parasitic diseases, with an average of 27.1 YPLL per death. As for males, the largest contributors to YPLL were deaths due to external causes of morbidity and mortality (18.3%; 18394), with an average of 37 YPLL per death, Table 5.

**Table 5:** Years of potential life lost by sex and cause of death, 2016 to 2020.

| Cause of death | Male |  |  | Female |  |  | Total |  |  |
| --- | --- | --- | --- | --- | --- | --- | --- | --- | --- |
|  | YPLL | % | YPLL per death | YPLL | % | YPLL per death | YPLL | % | YPLL per death |
| Certain conditions originating in the perinatal period | 142 | 0.1 | 71.0 | 71.0 | 0.2 | 71.0 | 213.0 | 0.1 | 71.0 |
| External causes of morbidity and mortality | 18393.5 | 18.3 | 37.0 | 2726.5 | 6.0 | 37.9 | 21120.0 | 14.5 | 37.1 |
| Diseases of the genitourinary system | 746.0 | 0.7 | 14.9 | 347.5 | 0.8 | 12.9 | 1093.5 | 0.8 | 14.2 |
| Diseases of the circulatory system | 15921.0 | 15.9 | 17.9 | 8881.5 | 19.7 | 16.5 | 24802.5 | 17.0 | 17.4 |
| Diseases of the digestive system | 5118.0 | 5.1 | 22.4 | 2159.0 | 4.8 | 24.5 | 7277.0 | 5.0 | 23.0 |
| Diseases of the respiratory system | 9793.0 | 9.8 | 22.5 | 4681.0 | 10.4 | 24.5 | 14474.0 | 9.9 | 23.1 |
| Mental and behavioral disorders | 3664.0 | 3.6 | 24.8 | 354.5 | 0.8 | 27.3 | 4018.5 | 2.8 | 25.0 |
| Endocrine, nutritional and metabolic diseases | 2455.5 | 2.4 | 23.2 | 1126.0 | 2.5 | 20.1 | 3581.5 | 2.5 | 22.1 |
| Diseases of the nervous system | 2603.0 | 2.6 | 31.4 | 1195.5 | 2.6 | 37.4 | 3798.5 | 2.6 | 33.0 |
| Pregnancy, childbirth and puerperium | NA* | NA* | NA* | 1183.5 | 2.6 | 40.8 | 1183.5 | 0.8 | 40.8 |
| Certain infectious and parasitic diseases | 14297.5 | 14.2 | 25.0 | 9984.0 | 22.1 | 27.1 | 24281.5 | 16.7 | 25.8 |
| Injury, poisoning and certain other consequences of external causes | 13632.0 | 13.6 | 32.9 | 2959.5 | 6.6 | 41.7 | 16591.5 | 11.4 | 34.2 |
| Congenital malformations, deformations and chromosomal abnormalities | 137.5 | 0.1 | 68.8 | 127.5 | 0.3 | 63.8 | 265.0 | 0.2 | 66.3 |
| Diseases of the musculoskeletal system and connective tissue | 84.5 | 0.1 | 28.2 | 51.5 | 0.1 | 51.5 | 136.0 | 0.1 | 34.0 |
| Diseases of the skin and subcutaneous tissue | 60.5 | 0.1 | 10.1 | 38.0 | 0.1 | 19.0 | 98.5 | 0.1 | 12.3 |
| Diseases of the blood and blood-forming organs and certain disorders involving the immune mechanism | 142.5 | 0.1 | 28.5 | 298.0 | 0.7 | 33.1 | 440.5 | 0.3 | 31.5 |
| Symptoms, signs and abnormal clinical and laboratory findings, not elsewhere classified | 3730.5 | 3.7 | 24.9 | 1897.0 | 4.2 | 25.3 | 5627.5 | 3.9 | 25.0 |
| Neoplasms [tumors] | 9468.0 | 9.4 | 16.8 | 7072.5 | 15.7 | 19.1 | 16540.5 | 11.4 | 17.7 |
| **Total** | **100389.0** | **100** | **24.2** | **45154.0** | **100** | **23.2** | **145543.0** | **100.0** | **23.9** |

Legend for Table. 5 \* NA – Not applicable

Overall, males contributed a higher number of YPLL throughout the study period (2016 – 2020). The highest proportion of YPLL (62.2%; 62479) occurred in the age group from 30 to 59 years for males. In the female population, the highest proportion of YPLL corresponded to range 1 to 4 (11.3%; 5112) followed by 35 to 39 age group (10.5%; 4745) (Fig 1).

**Figure 1.**
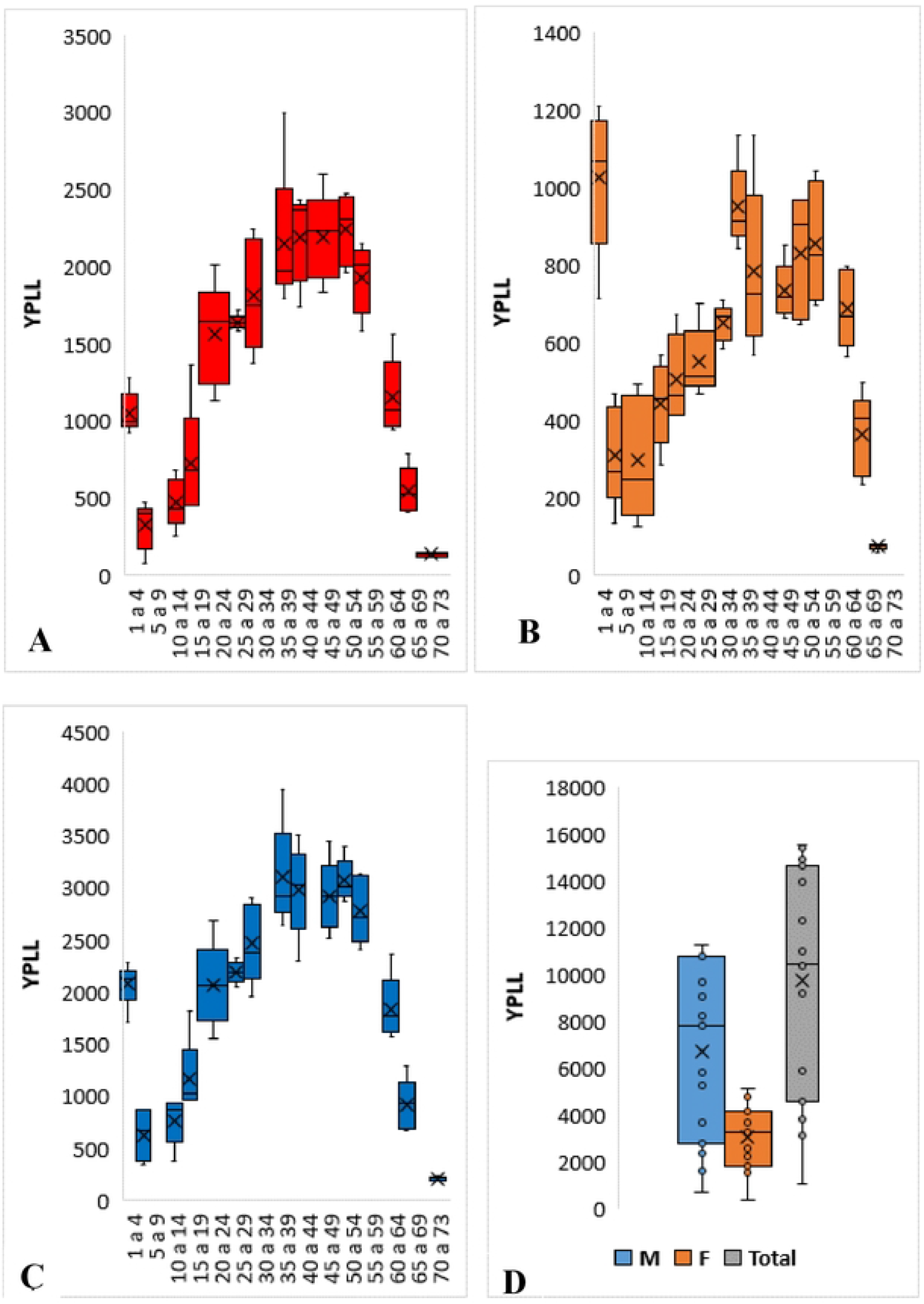
Distribution of Years of potential life lost by sex and age group, 2016 to 2020. Legend for-Fig 1. A is the distribution of YPLL for males, B is the distribution of YPLL for males and females, C and D represent the distribution of YPLL for both sexes.

### Estimation of years of potential productive life lost

From 2016 to 2020, there was a total loss of 80,965 YPPLL in Cabo Verde considering the 4,634 deaths occurred in the productive age group. There was a greater loss of years of productive life among men (72.1%; 58403) when compared to women (27.9%; 22563), configuring an annual average of 11680.8 YPPLL for men and 3926.2 for women, respectively, Table 6.

**Table 6:** Years of potential productive life lost by sex, 2016 to 2020.

| Year | Male |  |  | Female |  |  | Total |  |  |
| --- | --- | --- | --- | --- | --- | --- | --- | --- | --- |
|  | YPPLL | YPPLL rate | Mean | YPPLL | YPPLL rate | Mean | YPPLL | YPPLL rate | Mean |
| 2016 | 12095.0 | 68.1 | 19.3 | 4622.5 | 27.1 | 16.9 | 16717.5 | 48.0 | 18.6 |
| 2017 | 9847.5 | 54.4 | 17.0 | 4195.0 | 24.2 | 17.1 | 14042.5 | 39.6 | 17.0 |
| 2018 | 12565.0 | 68.1 | 18.2 | 4762.5 | 27.1 | 15.9 | 17327.5 | 48.1 | 17.5 |
| 2019 | 11502.5 | 61.2 | 18.0 | 4142.5 | 23.3 | 16.6 | 15645.0 | 42.8 | 17.6 |
| 2020 | 12392.5 | 64.8 | 17.0 | 4840.0 | 26.9 | 16.1 | 17232.5 | 46.5 | 16.7 |

Overall, the highest YPPLL values were recorded in 2018 and 2020, and YPPLL in the study period varied from 14,043 to 17,328, Table 7. In the 5-year study period, the largest contributors to YPPLL were external causes of morbidity and mortality, responsible for 17.9% (14,525) of total YPPLL, certain infectious and parasitic diseases 16.8% (13,583), diseases of the circulatory system 15.9% (12,878), injuries, poisoning and certain other consequences of external causes 11.2% (9,038), neoplasms [tumors] 10.6% (8,583) and diseases of the respiratory system 8.1% (6,528), Table 7.

**Table 7:**
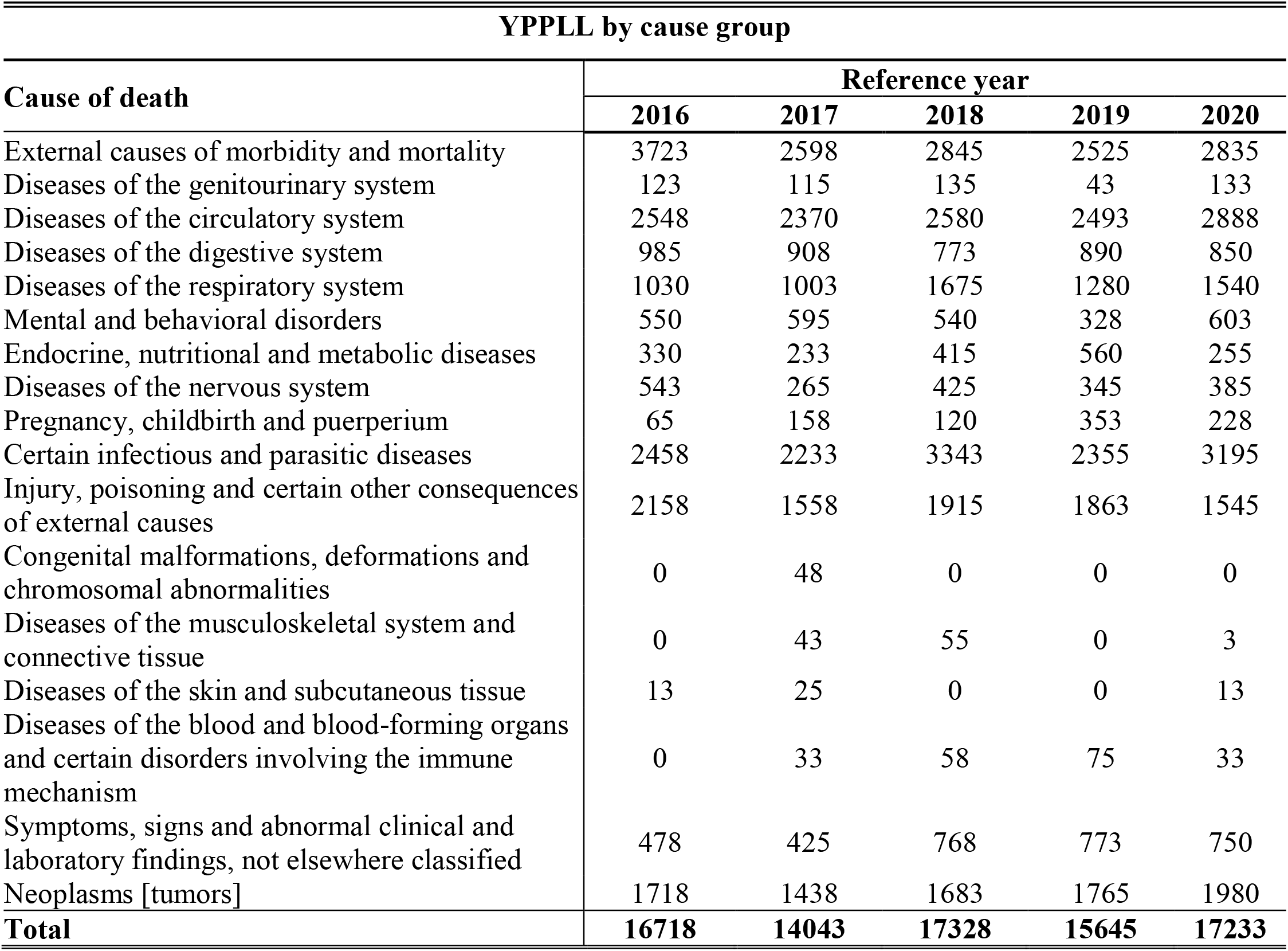
Evolution of years of potential productive life lost, 2016 to 2020.

| YPPLL by cause group |  |  |  |  |  |
| --- | --- | --- | --- | --- | --- |
| Cause of death | Reference year |  |  |  |  |
|  | 2016 | 2017 | 2018 | 2019 | 2020 |
| External causes of morbidity and mortality | 3723 | 2598 | 2845 | 2525 | 2835 |
| Diseases of the genitourinary system | 123 | 115 | 135 | 43 | 133 |
| Diseases of the circulatory system | 2548 | 2370 | 2580 | 2493 | 2888 |
| Diseases of the digestive system | 985 | 908 | 773 | 890 | 850 |
| Diseases of the respiratory system | 1030 | 1003 | 1675 | 1280 | 1540 |
| Mental and behavioral disorders | 550 | 595 | 540 | 328 | 603 |
| Endocrine, nutritional and metabolic diseases | 330 | 233 | 415 | 560 | 255 |
| Diseases of the nervous system | 543 | 265 | 425 | 345 | 385 |
| Pregnancy, childbirth and puerperium | 65 | 158 | 120 | 353 | 228 |
| Certain infectious and parasitic diseases | 2458 | 2233 | 3343 | 2355 | 3195 |
| Injury, poisoning and certain other consequences of external causes | 2158 | 1558 | 1915 | 1863 | 1545 |
| Congenital malformations, deformations and chromosomal abnormalities | 0 | 48 | 0 | 0 | 0 |
| Diseases of the musculoskeletal system and connective tissue | 0 | 43 | 55 | 0 | 3 |
| Diseases of the skin and subcutaneous tissue | 13 | 25 | 0 | 0 | 13 |
| Diseases of the blood and blood-forming organs and certain disorders involving the immune mechanism | 0 | 33 | 58 | 75 | 33 |
| Symptoms, signs and abnormal clinical and laboratory findings, not elsewhere classified | 478 | 425 | 768 | 773 | 750 |
| Neoplasms [tumors] | 1718 | 1438 | 1683 | 1765 | 1980 |
| **Total** | **16718** | **14043** | **17328** | **15645** | **17233** |

Males accounted for 72.1% (58,403) of YPPLL, with an average of 17.9 YPPLL per death, compared to 27.9% (22,563) for females, with a mean of 16.5 per death. Overall, external causes of morbidity and mortality were responsible for 14,525 (17.9%) of YPPLL with an average of 27.9 YPPLL per death. Results demonstrated that the major contributors to YPPLL for males were external causes of morbidity and mortality (22.0%; 12,843) with a mean of 28.1 per death in that specific population. As for the female population, certain infectious and parasitic diseases were the principal contributors (24.6%; 5,560), with an average of 18.7 YPPLL per death Table 8.

**Table 8:** Years of potential productive life lost by cause of death, 2016 to 2020.

| Cause of death | Male |  |  | Female |  |  | Total |  |  |
| --- | --- | --- | --- | --- | --- | --- | --- | --- | --- |
|  | YPPLL | % | Mean | YPPLL | % | Mean | YPPLL | % | Mean |
| External causes of morbidity and mortality | 12842.5 | 22.0 | 28.1 | 1682.5 | 7.5 | 26.7 | 14525 | 17.9 | 27.9 |
| Diseases of the genitourinary system | 397.5 | 0.7 | 12.8 | 150.0 | 0.7 | 9.4 | 547.5 | 0.7 | 11.6 |
| Diseases of the circulatory system | 8582.5 | 14.7 | 13.6 | 4295.0 | 19.0 | 12.9 | 12877.5 | 15.9 | 13.3 |
| Diseases of the digestive system | 3095.0 | 5.3 | 16.1 | 1310.0 | 5.8 | 18.7 | 4405 | 5.4 | 16.8 |
| Diseases of the respiratory system | 4747.5 | 8.1 | 15.4 | 1780.0 | 7.9 | 16.8 | 6527.5 | 8.1 | 15.7 |
| Mental and behavioral disorders | 2377.5 | 4.1 | 17.4 | 237.5 | 1.1 | 18.3 | 2615 | 3.2 | 17.4 |
| Endocrine, nutritional and metabolic diseases | 1285.0 | 2.2 | 16.5 | 507.5 | 2.2 | 13.0 | 1792.5 | 2.2 | 15.3 |
| Diseases of the nervous system | 1460.0 | 2.5 | 20.9 | 502.5 | 2.2 | 26.4 | 1962.5 | 2.4 | 22.1 |
| Pregnancy, childbirth and puerperium | 0.0 | 0.0 | 0.0 | 922.5 | 4.1 | 31.8 | 922.5 | 1.1 | 31.8 |
| Certain infectious and parasitic diseases | 8022.5 | 13.7 | 17.6 | 5560.0 | 24.6 | 18.7 | 13582.5 | 16.8 | 18.0 |
| Injury, poisoning and certain other consequences of external causes | 8135.0 | 13.9 | 22.7 | 902.5 | 4.0 | 21.0 | 9037.5 | 11.2 | 22.5 |
| Congenital malformations, deformations and chromosomal abnormalities | 0.0 | 0.0 | 0.0 | 47.5 | 0.2 | 47.5 | 47.5 | 0.1 | 47.5 |
| Diseases of the musculoskeletal system and connective tissue | 57.5 | 0.1 | 19.2 | 42.5 | 0.2 | 42.5 | 100 | 0.1 | 25.0 |
| Diseases of the skin and subcutaneous tissue | 27.5 | 0.0 | 9.2 | 22.5 | 0.1 | 22.5 | 50 | 0.1 | 12.5 |
| Diseases of the blood and blood-forming organs and certain disorders involving the immune mechanism | 97.5 | 0.2 | 19.5 | 100.0 | 0.4 | 16.7 | 197.5 | 0.2 | 18.0 |
| Symptoms, signs and abnormal clinical and laboratory findings, not elsewhere classified | 2340.0 | 4.0 | 20.2 | 852.5 | 3.8 | 17.4 | 3192.5 | 3.9 | 19.3 |
| Neoplasms [tumors] | 4935.0 | 8.4 | 11.8 | 3647.5 | 16.2 | 13.1 | 8582.5 | 10.6 | 12.3 |
| **Total** | **58402.5** | **100** | **17.9** | **22562.5** | **100** | **16.5** | **80965.0** | **100** | **17.5** |

The highest proportion of losses occurred in the 35-39 and 40-44 age groups and represented 27.3% and 28.2% of the total YPPLL, in the male and female population, respectively. The highest YPPLL recorded was in the range 35 to 39 years and corresponded to 2255 YPPLL while the lowest YPPLL was 205 in the 60 to 64 years age group, in the male population. For females, the range 35 to 39 registered the highest number of YPPLL corresponded to 852 and the minimum record of productivity losses, 123, was in the range 60 to 64 years. (Fig 2).

**Figure 2.**
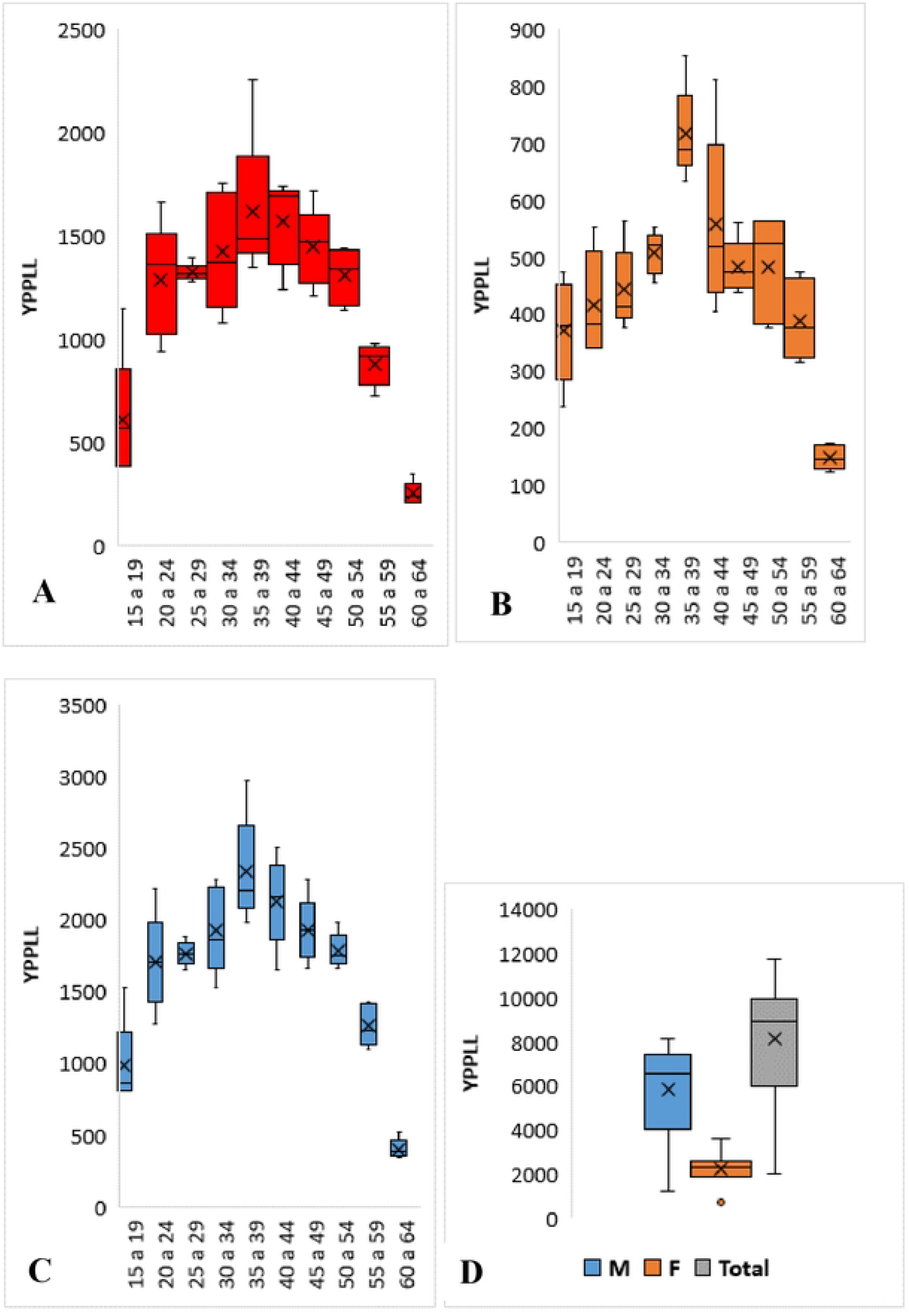
Distribution of Years of potential productive life lost by sex and age group, 2016 to 2020. Legend for Fig 1. A is the distribution of YPPLL for males, B represents the distribution of YPPLL for males and females, C and D – represent the distribution of YPPLL for both sexes.

### Economic impact of premature death

Premature mortality in the period from 2016 to 2020, resulted in a total of 98,659,153.23 USD of costs of productivity lost (CPL) with a mean of 21,290.28 USD per death. The CPL varied from 17,285,109.09 USD to 21,463,604.36 USD, while the leading contributors to CPL were circulatory system diseases (18,843,260.42 USD), external causes of morbidity and mortality (12,316,441.51 USD), certain infectious and parasitic diseases (16,633,842.70 USD), and neoplasms (13,580,684.86USD) and accounted for 62.2% of the total CPL. Tables 9 and 10.

**Table 9:** Cost of productivity lost (in USD) by cause of death, 2016 to 2020.

| Cause of death | 2016 | 2017 | 2018 | 2019 | 2020 |
| --- | --- | --- | --- | --- | --- |
| External causes of morbidity and mortality | 3073925.23 | 2183315.84 | 2415893.49 | 2194716.16 | 2448590.80 |
| Diseases of the genitourinary system | 167764.42 | 229093.17 | 256334.60 | 75154.97 | 168204.78 |
| Diseases of the circulatory system | 3564620.47 | 3539694.15 | 3808255.25 | 3561628.71 | 4369061.84 |
| Diseases of the digestive system | 1270545.30 | 1168387.24 | 1024750.91 | 1160451.01 | 1090518.36 |
| Diseases of the respiratory system | 1258969.32 | 1423126.38 | 2138614.40 | 1655213.04 | 1989788.77 |
| Mental and behavioral disorders | 736558.73 | 765967.70 | 764596.89 | 445149.45 | 793331.30 |
| Endocrine, nutritional and metabolic diseases | 522626.14 | 357363.09 | 592988.95 | 614479.74 | 410955.22 |
| Diseases of the nervous system | 598385.81 | 269465.21 | 387155.30 | 379429.36 | 414897.85 |
| Pregnancy, childbirth and puerperium | 51912.59 | 131210.84 | 107709.77 | 286243.07 | 180295.36 |
| Certain infectious and parasitic diseases | 2909533.92 | 2760117.89 | 3961300.28 | 3107916.68 | 3894973.93 |
| Injury, poisoning and certain other consequences of external causes | 2114697.85 | 1474031.68 | 1999671.55 | 2010407.09 | 1665704.75 |
| Congenital malformations, deformations and chromosomal abnormalities | 0.00 | 19796.71 | 0.00 | 0.00 | 0.00 |
| Diseases of the musculoskeletal system and connective tissue | 0.00 | 22073.43 | 51587.90 | 0.00 | 7552.18 |
| Diseases of the skin and subcutaneous tissue | 24315.30 | 35735.31 | 0.00 | 0.00 | 24315.30 |
| Diseases of the blood and blood-forming organs and certain disorders involving the immune mechanism | 0.00 | 26213.63 | 107040.73 | 47437.97 | 66811.36 |
| Symptoms, signs and abnormal clinical and laboratory findings, not elsewhere classified | 626656.88 | 478989.03 | 788430.13 | 847065.51 | 958750.44 |
| Neoplasms [tumors] | 2726208.00 | 2400527.81 | 2784972.24 | 2689124.69 | 2979852.12 |
| **Total** | **19646719.95** | **17285109.09** | **21189302.39** | **19074417.45** | **21463604.36** |

**Table 10:**
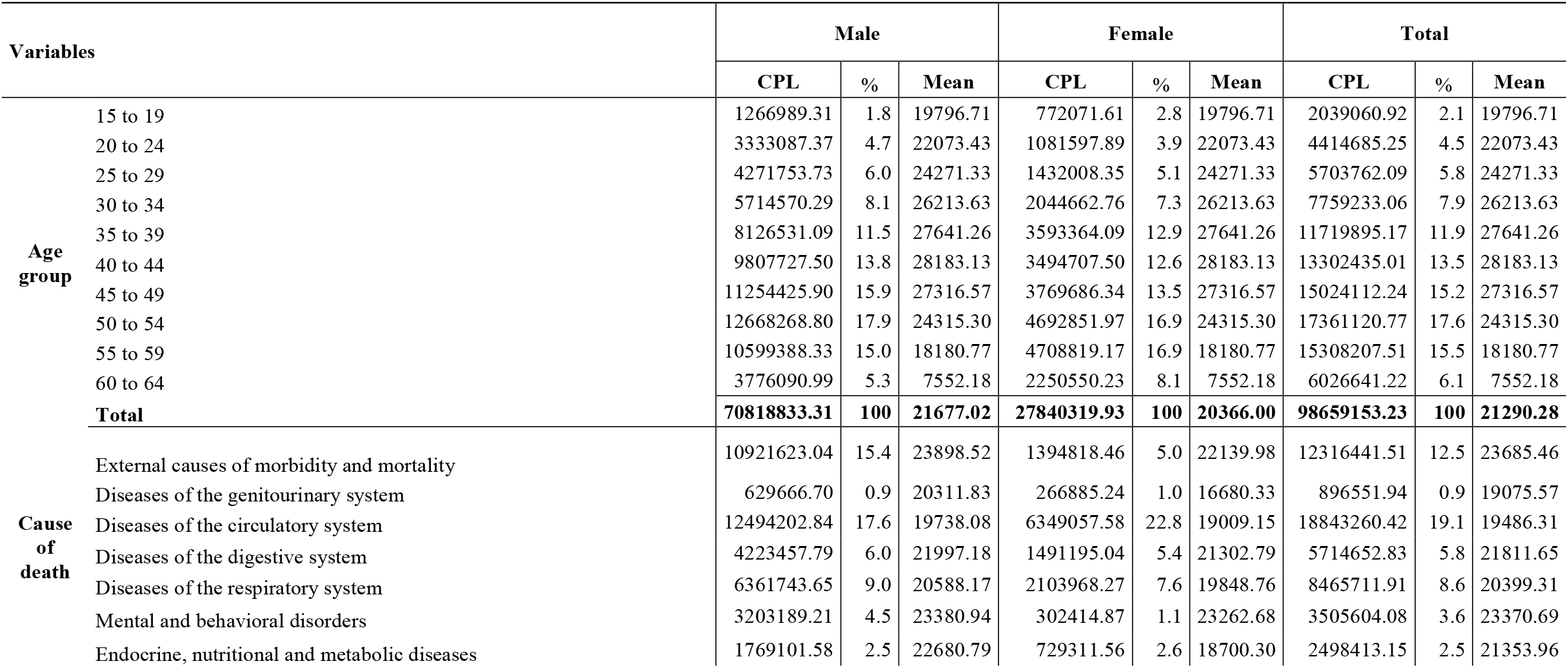
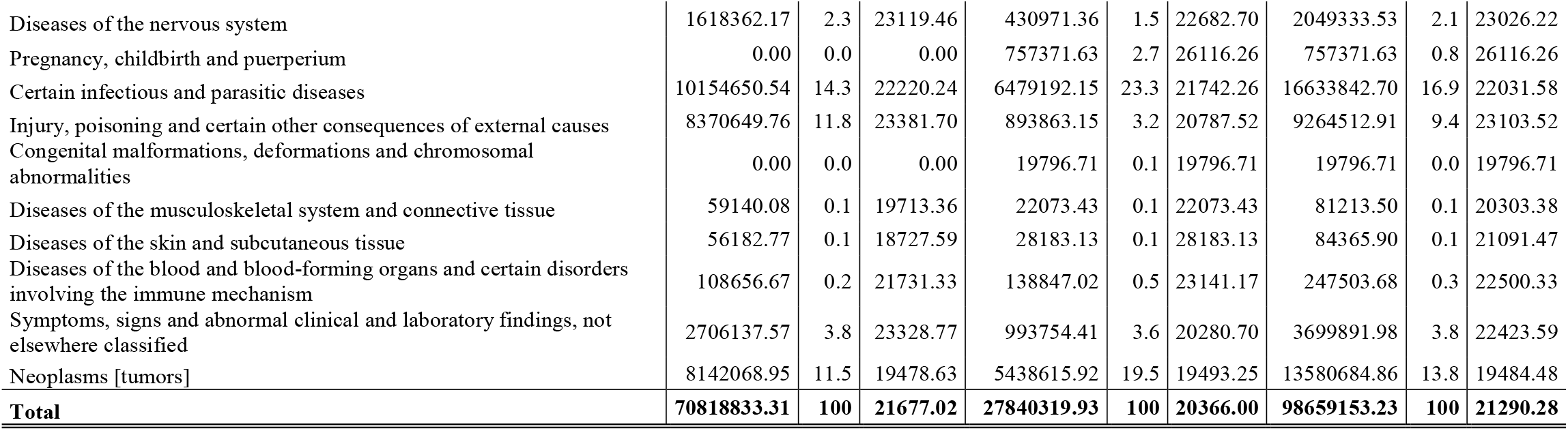
Cost of productivity lost by sex, age group, and cause of death, 2016 to 2020.

| Variables |  | Male |  |  | Female |  |  | Total |  |  |
| --- | --- | --- | --- | --- | --- | --- | --- | --- | --- | --- |
|  |  | CPL | % | Mean | CPL | % | Mean | CPL | % | Mean |
| Age group | 15 to 19 | 1266989.31 | 1.8 | 19796.71 | 772071.61 | 2.8 | 19796.71 | 2039060.92 | 2.1 | 19796.71 |
|  | 20 to 24 | 3333087.37 | 4.7 | 22073.43 | 1081597.89 | 3.9 | 22073.43 | 4414685.25 | 4.5 | 22073.43 |
|  | 25 to 29 | 4271753.73 | 6.0 | 24271.33 | 1432008.35 | 5.1 | 24271.33 | 5703762.09 | 5.8 | 24271.33 |
|  | 30 to 34 | 5714570.29 | 8.1 | 26213.63 | 2044662.76 | 7.3 | 26213.63 | 7759233.06 | 7.9 | 26213.63 |
|  | 35 to 39 | 8126531.09 | 11.5 | 27641.26 | 3593364.09 | 12.9 | 27641.26 | 11719895.17 | 11.9 | 27641.26 |
|  | 40 to 44 | 9807727.50 | 13.8 | 28183.13 | 3494707.50 | 12.6 | 28183.13 | 13302435.01 | 13.5 | 28183.13 |
|  | 45 to 49 | 11254425.90 | 15.9 | 27316.57 | 3769686.34 | 13.5 | 27316.57 | 15024112.24 | 15.2 | 27316.57 |
|  | 50 to 54 | 12668268.80 | 17.9 | 24315.30 | 4692851.97 | 16.9 | 24315.30 | 17361120.77 | 17.6 | 24315.30 |
|  | 55 to 59 | 10599388.33 | 15.0 | 18180.77 | 4708819.17 | 16.9 | 18180.77 | 15308207.51 | 15.5 | 18180.77 |
|  | 60 to 64 | 3776090.99 | 5.3 | 7552.18 | 2250550.23 | 8.1 | 7552.18 | 6026641.22 | 6.1 | 7552.18 |
|  | **Total** | **70818833.31** | **100** | **21677.02** | **27840319.93** | **100** | **20366.00** | **98659153.23** | **100** | **21290.28** |
| Cause of death | External causes of morbidity and mortality | 10921623.04 | 15.4 | 23898.52 | 1394818.46 | 5.0 | 22139.98 | 12316441.51 | 12.5 | 23685.46 |
|  | Diseases of the genitourinary system | 629666.70 | 0.9 | 20311.83 | 266885.24 | 1.0 | 16680.33 | 896551.94 | 0.9 | 19075.57 |
|  | Diseases of the circulatory system | 12494202.84 | 17.6 | 19738.08 | 6349057.58 | 22.8 | 19009.15 | 18843260.42 | 19.1 | 19486.31 |
|  | Diseases of the digestive system | 4223457.79 | 6.0 | 21997.18 | 1491195.04 | 5.4 | 21302.79 | 5714652.83 | 5.8 | 21811.65 |
|  | Diseases of the respiratory system | 6361743.65 | 9.0 | 20588.17 | 2103968.27 | 7.6 | 19848.76 | 8465711.91 | 8.6 | 20399.31 |
|  | Mental and behavioral disorders | 3203189.21 | 4.5 | 23380.94 | 302414.87 | 1.1 | 23262.68 | 3505604.08 | 3.6 | 23370.69 |
|  | Endocrine, nutritional and metabolic diseases | 1769101.58 | 2.5 | 22680.79 | 729311.56 | 2.6 | 18700.30 | 2498413.15 | 2.5 | 21353.96 |

|  |  |  |  |  |  |  |  |  |  |
| --- | --- | --- | --- | --- | --- | --- | --- | --- | --- |
| Diseases of the nervous system | 1618362.17 | 2.3 | 23119.46 | 430971.36 | 1.5 | 22682.70 | 2049333.53 | 2.1 | 23026.22 |
| Pregnancy, childbirth and puerperium | 0.00 | 0.0 | 0.00 | 757371.63 | 2.7 | 26116.26 | 757371.63 | 0.8 | 26116.26 |
| Certain infectious and parasitic diseases | 10154650.54 | 14.3 | 22220.24 | 6479192.15 | 23.3 | 21742.26 | 16633842.70 | 16.9 | 22031.58 |
| Injury, poisoning and certain other consequences of external causes | 8370649.76 | 11.8 | 23381.70 | 893863.15 | 3.2 | 20787.52 | 9264512.91 | 9.4 | 23103.52 |
| Congenital malformations, deformations and chromosomal abnormalities | 0.00 | 0.0 | 0.00 | 19796.71 | 0.1 | 19796.71 | 19796.71 | 0.0 | 19796.71 |
| Diseases of the musculoskeletal system and connective tissue | 59140.08 | 0.1 | 19713.36 | 22073.43 | 0.1 | 22073.43 | 81213.50 | 0.1 | 20303.38 |
| Diseases of the skin and subcutaneous tissue | 56182.77 | 0.1 | 18727.59 | 28183.13 | 0.1 | 28183.13 | 84365.90 | 0.1 | 21091.47 |
| Diseases of the blood and blood-forming organs and certain disorders involving the immune mechanism | 108656.67 | 0.2 | 21731.33 | 138847.02 | 0.5 | 23141.17 | 247503.68 | 0.3 | 22500.33 |
| Symptoms, signs and abnormal clinical and laboratory findings, not elsewhere classified | 2706137.57 | 3.8 | 23328.77 | 993754.41 | 3.6 | 20280.70 | 3699891.98 | 3.8 | 22423.59 |
| Neoplasms [tumors] | 8142068.95 | 11.5 | 19478.63 | 5438615.92 | 19.5 | 19493.25 | 13580684.86 | 13.8 | 19484.48 |
| **Total** | **70818833.31** | **100** | **21677.02** | **27840319.93** | **100** | **20366.00** | **98659153.23** | **100** | **21290.28** |

Males accounted for 71.8% (70,818,833.31 USD) with a mean of 21,677.02 USD per death while females were responsible for 28.2% (27,840,319.93 USD) of the total CPL, and had an average of 20,366.00 USD per death, respectively. In general, the highest CPLs were recorded in the age groups between 35 and 59 years (73.7%; 72,715,770.69 USD) Table 10. Analysis showed that Pregnancy, childbirth and puerperium had the highest mean CPL per death of 26,116.26 USD, followed by the “external causes of morbidity and mortality” group, with an average CPL of US$ 23,685.46 and Mental and behavioral disorders with a mean of 23,370.69 USD per death, Table 10**: Cost of productivity lost by sex, age group, and cause**.

The sensitivity analysis was performed and showed total CPL of 134,724,671.62 USD (mean = 29 073.08 USD /death) at a discount rate of 3% and 74,517,897.02 USD (mean = 16,080.69 USD /death) at 6%. Diseases of the circulatory system had the highest CPL observed at a rate of 3%, and accounted for 24,297,098.13 USD or 18.0% of total CPL, followed by certain infectious and parasitic diseases with 22,809,540.03 USD (16.9%). At a rate of 6%, the same trend was observed, with the two main causes of death having highest CPL, Table 11.

**Table 11:** Lost productivity cost using GDP per capita at 3% and 6% discount rate, 2016 to 2020.

| Cause of death | Present value (Total) |  |  |  |  |  |
| --- | --- | --- | --- | --- | --- | --- |
|  | 3% | % | Mean | 6% | % | Mean |
| External causes of morbidity and mortality | 19015041.77 | 14.1 | 36567.39 | 8250513.82 | 11.1 | 15866.37 |
| Diseases of the genitourinary system | 1122230.56 | 0.8 | 23877.25 | 729652.17 | 1.0 | 15524.51 |
| Diseases of the circulatory system | 24297098.13 | 18.0 | 25126.26 | 14990278.74 | 20.1 | 15501.84 |
| Diseases of the digestive system | 7694029.08 | 5.7 | 29366.52 | 4352584.69 | 5.8 | 16612.92 |
| Diseases of the respiratory system | 11334604.38 | 8.4 | 27312.30 | 6512545.57 | 8.7 | 15692.88 |
| Mental and behavioral disorders | 4703578.15 | 3.5 | 31357.19 | 2664327.91 | 3.6 | 17762.19 |
| Endocrine, nutritional and metabolic diseases | 3284790.53 | 2.4 | 28075.13 | 1946388.86 | 2.6 | 16635.80 |
| Diseases of the nervous system | 2939208.04 | 2.2 | 33024.81 | 1479318.09 | 2.0 | 16621.55 |
| Pregnancy, childbirth and puerperium | 1204568.78 | 0.9 | 41536.85 | 482418.45 | 0.6 | 16635.12 |
| Certain infectious and parasitic diseases | 22809540.03 | 16.9 | 30211.31 | 12456681.11 | 16.7 | 16498.92 |
| Injury, poisoning and certain other consequences of external causes | 13427871.31 | 10.0 | 33485.96 | 6598101.86 | 8.9 | 16454.12 |
| Congenital malformations, deformations and chromosomal abnormalities | 39626.72 | 0.0 | 39626.72 | 9988.34 | 0.0 | 9988.34 |
| Diseases of the musculoskeletal system and connective tissue | 126469.38 | 0.1 | 31617.35 | 54453.07 | 0.1 | 13613.27 |
| Diseases of the skin and subcutaneous tissue | 105874.72 | 0.1 | 26468.68 | 67948.23 | 0.1 | 16987.06 |
| Diseases of the blood and blood-forming organs and certain disorders involving the immune mechanism | 332274.62 | 0.2 | 30206.78 | 190714.50 | 0.3 | 17337.68 |
| Symptoms, signs and abnormal clinical and laboratory findings, not elsewhere classified | 5151758.37 | 3.8 | 31222.78 | 2737935.51 | 3.7 | 16593.55 |
| Neoplasms [tumors] | 17136107.06 | 12.7 | 24585.52 | 10994046.11 | 14.8 | 15773.38 |
| **Total** | **134724671.62** | **100** | **29073.08** | **74517897.02** | **100** | **16080.69** |

## Discussion of results

The present study estimated the loss of productivity due to premature mortality in Cabo Verde, from 2016 to 2020. For this, the Human Capital approach method was used in the analysis of the cost of the disease and the productivity losses due to premature mortality,[4,18]. The current study is the first conducted in Cabo Verde, assessing the indirect costs due to all-causes of premature mortality.

The results of the present study point to a significant difference between the sexes, in terms of productivity losses. Males accounted for 71.8% of the total CPL. This result was higher than the results obtained in Tanzania and Iran, where the costs attributed to men were around 58% and 67%, respectively,[4.18]. However, it is noteworthy that in the last two studies, the authors evaluated only the main causes of death, while in the present study, all causes of premature death were taken into account,[4,18]. Furthermore, the Łyszczarz [19] reported costs attributed to males between 64.7% and 81.2% of the total costs, in Europe, while Díaz-Jiménez et al.[20] revealed that the CPL for males was three times higher for females. The predominance of males shown in the current analysis reflected the burden of premature mortality in this population. In other economic studies conducted by Menzin et al. and Carter et al.[17,21], the notable burden of lost productivity attributed to males was associated with greater participation in economic activities and disparity in income between the sexes.

In the present study, the estimated 8,965 YPPLL calculated resulted in a total loss of 98,659,153.23 USD and an average 21,290.28 USD per death. Total estimated CPL represented 5.3% of GDP (1,853,485,384.00 USD) in 2017,[14]. The value of CPL in relation to the GDP in the current study was lower than that reported by Łyszczarz [19],which showed potential losses between 0.6% and 3.2% of GDP in the evaluated European countries. Díaz-Jiménez et al. [20] reported values between 1.6 and 3.0% while Rumisha et al. [4] reported values equivalent to 0.3% of GDP.

In the present study, the mean CPL per death was 21,290.28 USD, a value higher than the results of Rumisha et al.[4], and lower than the mean value per death reported by Najafi et al.[18]. In the current analysis, the group “pregnancy, childbirth and puerperium” represented the highest CPL average per death (26,116.26 USD) and only 0.8% of the estimated total CPL. In comparison, this group accounted for a total loss of productivity of less than 0.1%, in a similar study [21] and 1% of lost productivity in the African region [22].

When mortality data were analyzed from a productivity loss perspective, results showed that the main drivers of CPL were trauma, external causes of morbidity and mortality (12.5%) and Injury, poisoning and certain other consequences of external causes (9.4%) represented 21.9% of the total CPL. Diseases of the circulatory system accounted for (19.1%), certain infectious and parasitic diseases 16.7% and neoplasms 13.8%. When comparing with other studies evaluating costs of productivity loss from all causes of premature death, Carter et al. [21] reported neoplasms (30%) as the biggest driver of lost productivity, followed by cardiovascular diseases (19%), unintentional injuries (15%) and intentional injuries (13%). Rumisha et al. [4] reported the predominance of communicable diseases, followed by neoplasms and injuries among the main contributors to CPL, and Diaz-Jimenez et al. [20] showed that injuries were responsible for 61% of lost productivity in their study.

The 921 deaths verified related to trauma in this study, occurred mostly in the young population and contributed to about 23562.5 YPPLL, which translated in 21,580,954.42 USD. For this cause of death, the age groups of 20 to 44 years represented 78% of YPPLL. In comparison, the 967 deaths due to diseases of the circulatory system, which resulted in 12,877.5 YPPLL approximately 50% less than YPPLL due to trauma. Showing that death due to trauma occurred in younger age groups. Diseases of the circulatory system corresponded to a CPL of 18,843,260.42 USD, and these deaths occurred mainly in the older age groups (age group 40 to 59 years represented 59.4% of YPPLL due to this cause). The results of the present study corroborate the findings by Delgado and Carter et al. [7,21] that demonstrated that death due to injuries often occur in the young population.

The results of the present study showed that the highest productivity loss was due to non-communicable diseases. For example, diseases of the circulatory system and neoplasms represented 32.9% of the total CPL. This finding is in line with the results of studies carried out in Iran [18] and in Australia [21], but differed from results in Tanzania [4] which reported a predominance of communicable diseases. However, communicable diseases continue to represent an important burden in the mortality profile of Cabo Verde.

For instance, certain infectious and parasitic diseases accounted for 16.7% of CPL, showing that the country is still in epidemiological transition [23]. Interestingly, this cause of death was the main driver of costs in the female population, accounting for 23.3% of the total CPL in this group, while diseases of the circulatory system and neoplasms contributed with 22.8% and 19.5%, respectively. Evidence available on the economic burden of disease on the African Continent [22] indicated a near balance between non-communicable diseases (37%) and communicable diseases (36%) as the main drivers of productivity losses, and this represents a challenge for health systems that have traditionally focused on communicable diseases [24]. It also emphasizes the need for greater investment in health and demonstrates the need to implement robust public health policies that address these issues [22].

Sensitivity analysis was performed according to the guidelines [25,26] for economic studies. The discount rates of 3% and 6% were applied, which resulted in present values of lost productivity of 134,724,671.62 USD and 74,517,897.02 USD, respectively. These values correspond to 7.3% and 4.0% of 2017 GDP [14].

There is a limited number of studies that assess productivity loss due to all causes of death, particularly on the African continent. In addition, most studies found analyzed costs due to specific causes [17,27–33]. Furthermore, no studies were found that addressed the issue of productivity loss in small island developing countries, such as Cabo Verde. Thus, the present study will contribute to the construction of scientific knowledge on the subject at national and regional level.

The present study had limitations that should be taken into account when interpreting the results. Firstly, it was assumed that each individual of working age would contribute equally to society if they did not die by the defined age limit using the Human Capital approach. Although this method is often criticized as it tends to overestimate the losses resulting from premature mortality [12,17,34], however, the method is widely used in economic studies, due to its relatively easy application. Moreover, there is still no consensus on the best methodology to assess the indirect costs of mortality [5,12,35]. Future economic studies evaluating the costs of premature death in Cabo Verde, could apply other methods described in the literature [17,19,21].

Other limitations include that fact that other indirect productivity losses related to mortality were not considered in the analysis, such as intangible costs, among others, which could lead to underestimation of the economic burden of the analyzed causes. Cost analysis was performed by group of causes. Therefore, the loss of productivity due to specific deaths was not demonstrated. This may constitute a topic for future investigations.

Finally, it was found that 4% of the calculated costs were attributed to the group of “symptoms, signs and abnormal findings of clinical and laboratory tests, not classified elsewhere”, which highlights the need to further improve the classification of causes of death and registration in the country’s health information system (SIS).

## Conclusion

The study demonstrated the burden of premature mortality and associated costs in Cabo Verde from 2016 to 2020 and elucidated patterns of early mortality in the period analyzed. This study revealed that the main drivers of lost productivity in the period were trauma, diseases of the circulatory system, certain infectious and parasitic diseases and neoplasms. It also showed that a significant proportion of the costs of lost productivity was attributable to males. Communicable diseases still represent an important burden with regard to the costs of lost productivity in Cabo Verde, showing that the country is still in an epidemiological transition.

The estimation of the burden of mortality and the cost of productivity lost for society and the national economy can be another instrument for assessing the mortality profile of the country, thus complementing the measures traditionally used to demonstrate the profile of mortality and support decision-making and allocation of resources. The implementation of policies aimed at reducing premature mortality, especially from preventable causes, may result in reduced costs of lost productivity and improvements in the well-being of the cabo-verdean population.

## Data Availability

The data underlying the results presented in the study are available from the National Directorate of Health, Ministry of Health, Cabo Verde

## Acknowledgments

The authors would like to thank the staff in charge of compiling mortality data and the National Health Directorate for authorizing the use of data.

## Author Contributions

**Conceptualization:** Ngibo Mubeta Fernandes, Edna Duarte Lopes, Janilza Solange Gomes Silveira Silva

**Data curation:** Janice de Jesus Xavier Soares, Domingos Veiga Varela

**Formal analysis**: Ngibo Mubeta Fernandes, Janice de Jesus Xavier Soares.

**Investigation:** Ngibo Mubeta Fernandes, Janice de Jesus Xavier Soares, Edna Duarte Lopes, Janilza Solange Gomes Silveira Silva

**Methodology**: Ngibo Mubeta Fernandes, Janice de Jesus Xavier Soares

**Project administration:** Edna Duarte Lopes

**Supervision:** Edna Duarte Lopes

**Visualization:** Ngibo Mubeta Fernandes, Janice de Jesus Xavier Soares

**Writing – original draft:** Ngibo Mubeta Fernandes, Janice de Jesus Xavier Soares, Janilza Solange Gomes Silveira Silva, Domingos Veiga Varela

**Writing – review & editing:** Ngibo Mubeta Fernandes, Edna Duarte Lopes, Janilza Solange Gomes Silveira Silva

## Supporting information

S1 Fig 1. Distribution of years of potential life lost by sex and age group, 2016 to 2020. Legend for Fig 1. A is the distribution of YPLL for males; B is the distribution of YPLL for females, C and D represent the distribution of YPLL for both sexes.

S2 Fig 2. Distribution of years of potential productive life lost by sex and age group, 2016 to 2020. Legend for Fig 2. A is the distribution of YPPLL for males; B is the distribution of YPPLL for females, C and D represent the distribution of YPPLL for both sexes.

S12 Annex 1: Characterization of deaths from all causes, Cabo Verde, 2016 to 2020 S13 Annex 2: Causes of death in the population, Cabo Verde, 2016 to 2020

S14 Annex 3: Rates of years of potential life lost by municipality, Cabo Verde, 2016 to 2020

S15 Annex 4: Rates of years of potential productive life lost by municipality, Cabo Verde, 2016 to 2020

